# Exercising caution in correlating COVID-19 incidence and mortality rates with BCG vaccination policies due to variable rates of SARS CoV-2 testing

**DOI:** 10.1101/2020.04.08.20056051

**Authors:** Janine Hensel, Kathleen M. McAndrews, Daniel J. McGrail, Dara P. Dowlatshahi, Valerie S. LeBleu, Raghu Kalluri

## Abstract

The Bacillus Calmette-Guerin (BCG) vaccine provides protection against tuberculosis (TB), and is proposed to provide protection to non-TB infectious diseases. The COVID-19 outbreak results from infection with the novel coronavirus SARS-CoV-2 (CoV-2) and was declared a pandemic on March 11^th^, 2020. We queried whether the BCG vaccine offers protection against CoV-2 infection. We observed that countries with a current universal BCG vaccination policy have a significantly lower COVID-19 incidence than countries which never had a universal BCG policy or had one in the past. However, population density, median age, TB incidence, urban population, and, most significantly, CoV-2 testing rate, were also connected with BCG policy and could potentially confound the analysis. By limiting the analysis to countries with high CoV-2 testing rates, defined as greater than 2,500 tests per million inhabitants, these parameters were no longer statistically associated with BCG policy. When analyzing only countries with high testing rates, there was no longer a significant association between the number of COVID-19 cases per million inhabitants and the BCG vaccination policy. Although preliminary, our analyses indicate that the BCG vaccination may not offer protection against CoV-2 infection. While reporting biases may confound our observations, our findings support exercising caution in determining potential correlation between BCG vaccination and COVID-19 incidence, in part due significantly lower rates of CoV-2 testing per million inhabitants in countries with current universal BCG vaccination policy.

## INTRODUCTION

Since the early 1920s, the use of the Bacillus Calmette-Guerin (BCG) vaccine has been implemented in several countries to prevent tuberculosis (TB), a disease caused by *Mycobacterium tuberculosis*. Infection with *Mycobacterium tuberculosis* primarily impacts lung function, and may lead to TB-related death. The vaccine consists in exposure to the *M. tuberculosis*-related, live, attenuated *Mycobacterium bovis* strain. The BCG vaccine thus offers a partial and possibly variable immunity against TB, and its off-target effect has also been linked to protection against other infectious diseases [1]. There is evidence from previous outbreak that supports the hypothesis that BCG vaccination can protect against RNA and DNA viral infections (herpes and influenza) and other disease such as asthma [2, 3]. These observations have given support to the possibility of the BCG vaccine providing protection against infection with the novel coronavirus, SARS CoV-2 (CoV-2). CoV-2 infection results in the disease known as COVID-19, which is associated with fever and severe respiratory illness. The WHO declared COVID-19 a pandemic on March 11^th^ 2020. As of April 7^th^, 2020, 81 countries around the globe reported over 500 COVID-19 cases.

In most countries with high TB incidence, children receive the BCG vaccine in early infancy. These countries have implemented a universal BCG vaccination policy. In some of these countries, when TB incidence dropped, the policy was amended to stop universal BCG vaccination, and this group is categorized in this report as having universal BCG policy in the past. A few countries have never had a universal BCG policy. The reason for not vaccinating was due to low TB incidence and the possibility to retain use of the tuberculin skin test in order to assess current TB infections. In countries with active BCG vaccination programs, the variability in protection against TB could be due to differences in the BCG strains used in vaccine synthesis, vaccine batch differences, time of vaccination, previous exposure to mycobacterium, and distinct host haplotypes [4]. The BCG vaccine may offer protection for as long as 30 to 40 years post vaccination (as previously reported in Norway [5]), and possibly as long as 50 to 60 years, as reported in American Indians and Alaska Natives that participated in a BCG vaccine trial [6]. In consideration for the potential protection of BCG vaccination against other infectious diseases including CoV-2 infection, Australia, which discontinued its universal BCG vaccination in the 1980s, is currently testing whether the BCG vaccination can protect healthcare workers from CoV-2 infection or reduce COVID-19-related symptoms (clinicaltrials.gov, NCT04327206). In this study, we investigated whether the BCG vaccine correlates with reported COVID-19 cases and mortality.

## METHODS

We collected information for BCG vaccination policies, BCG strains, and TB incidence across all countries from the BCG World Atlas (http://www.bcgatlas.org/). For analysis of countries with past BCG vaccination policy, countries with a year BCG was introduced and stopped were included. Data for Israel showed a change in policy in 1982, but did not specify cessation, therefore it was excluded from analysis. The BCG World Atlas was last updated in 2017. Data of total COVID-19 cases, new COVID-19 cases, first reported COVID-19 cases, mortality, and testing rates for each country was obtained April 7^th^, 2020 using https://www.worldometers.info/coronavirus/. Total population, population density (population/km^2^), net migrants, and median age were obtained from: https://www.worldometers.info/world-population/population-by-country/ (2020). Percent female population (2017) was obtained from: https://ourworldindata.org/grapher/share-population-female. Rate of smokers (2016) was obtained from: https://ourworldindata.org/smoking. Heart disease-related mortality (2002) was obtained from: https://www.who.int/cardiovascular_diseases/en/cvd_atlas_29_world_data_table.pdf World maps were generated using: https://mapchart.net/world.html.

Data were analyzed using IBM SPSS Statistics software (build 1.0.1.1347) and Matlab R2016a. In SPSS, BCG policy was used as the fixed variable, whereas “total COVID-19 cases per 1M population” and “percent mortality” defined as deaths per total COVID-19 cases were classified as dependent variables. Multivariate analysis was used to derive estimates of effect size and to compute homogeneity tests. Estimated marginal means were calculated and confidence interval adjustment was done using LSD post-hoc comparisons.

Univariate and multivariate regression analysis was performed using Matlab R2016a on log-transformed “total COVID-19 cases per 1M population” and “percent mortality” defined as deaths per total COVID-19 cases using the fitglme function. All confounding variables were z-normalized for equivalent scaling, and p values corrected using Benjamini and Hochberg procedure for univariate comparisons. BCG vaccination (BCG Vac.) was treated as continuous variable with 0 for never universal BCG policy, 1 for universal BCG policy in the past, and 2 for current universal BCG policy. For some comparisons, countries were limited to those with testing rates of 2,500 tests per million and greater, as specified in the results.

## RESULTS

As of April 7^th^, 2020, 211 countries around the world had reported COVID-19 cases. Among those, 81 countries had more than 500 confirmed COVID-19 cases. Using a 500 confirmed COVID-19 cases cut-off, we ascertained whether COVID-19 incidence and associated mortality is correlated with BCG vaccination policies. Three countries (Bahrain, Serbia, and Iceland) did not have information on vaccination policies. Out of the remaining 78 countries (**Table 1**), 8% of the countries never had a universal BCG policy (‘Never universal GCG policy’, **Figure 1A**). Belgium, Netherlands, Canada, Italy, Lebanon, and United States of America currently never had a universal BCG policy, yet all these countries may offer vaccination for high risk subgroups (healthcare professionals and children with parents from high risk countries). 23% of the analyzed countries had a universal BCG policy in the past, including many European countries, Australia, Ecuador, and Israel (‘Universal BCG policy in the past’, **Figure 1A**). 69% of the countries currently have a universal BCG vaccination policy, including most countries in Central America, South America, Africa, and Asia (‘Current universal BCG policy’, **Figure 1A**). The number of total COVID-19 cases (with a 500 cut-off) in each of the three groups is shown in **Figure 1B**. The number COVID-19 cases per 1 million (M) inhabitants was significantly reduced in countries with current universal BCG policy when compared to countries with a never- or in the past-universal BCG policy (**Figure 1C and D**). The percent mortality per total COVID-19 cases was significantly reduced in countries with current and past universal BCG policy when compared to countries that never had a universal BCG policy (**Figure 1E and F**). These analyses however indicated that other factors may confound the results, and included population density, median age, TB incidence, testing rate, and urban population. The following analyses addressed these potential confounders.

**Table 1.**
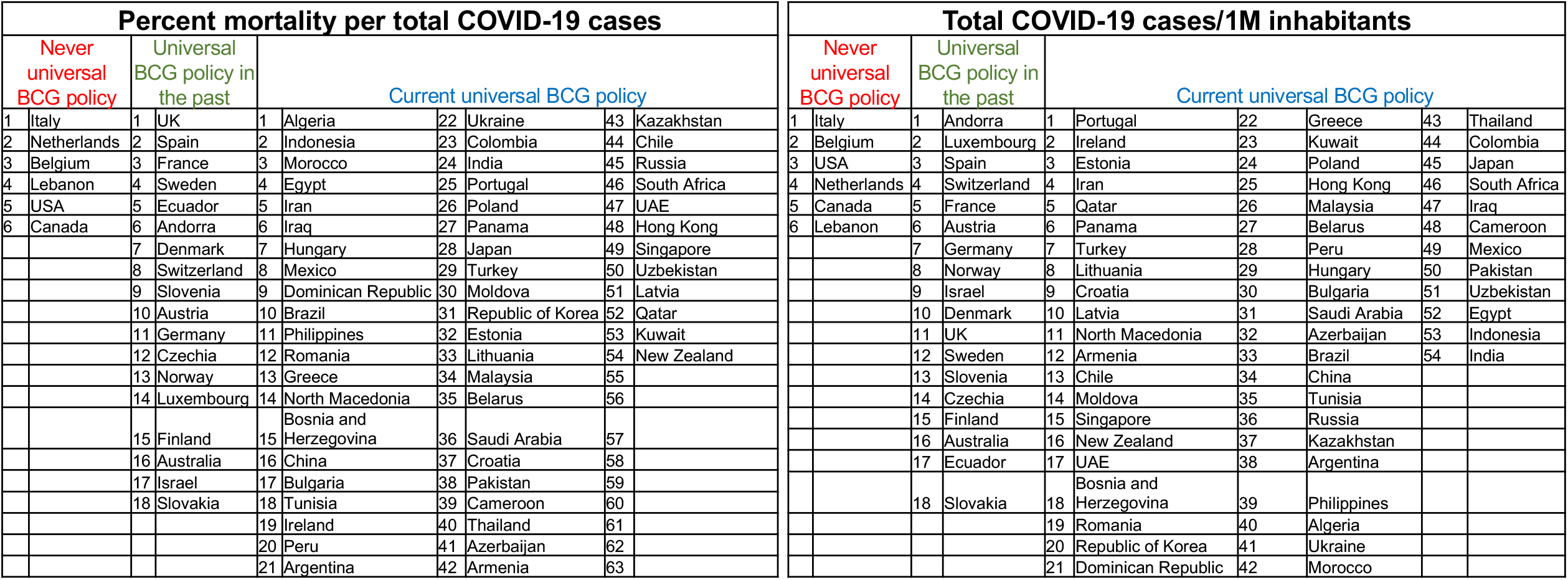
Countries are ranked from highest to lowest based on percent mortality per total COVID-19 cases and total COVID-19 cases /1M inhabitants.

**Figure 1.**
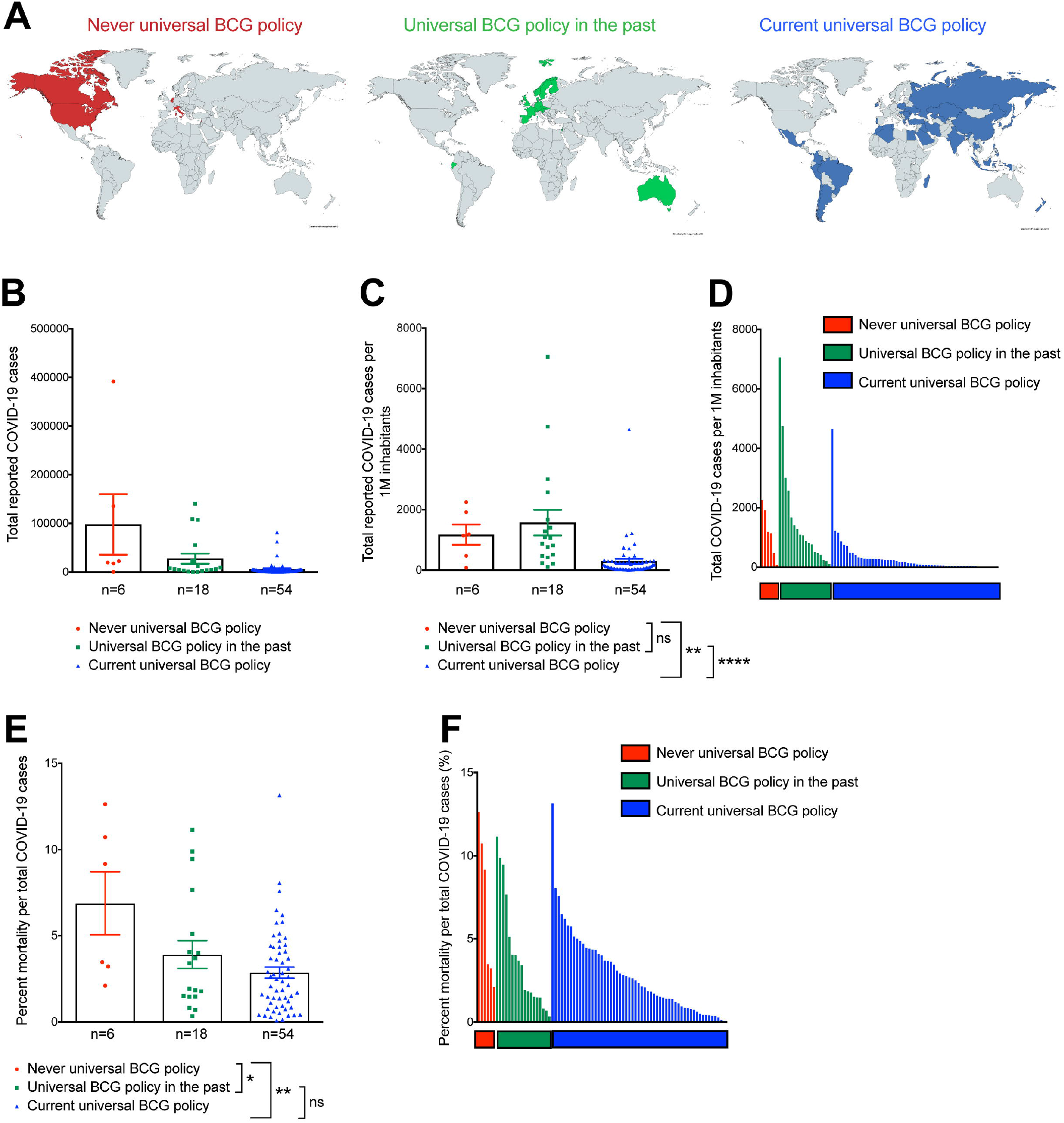
COVID-19 incidence and mortality in countries with distinct universal BCG vaccination policy. **(A)** World maps show countries that never had a universal BCG vaccination policy (in red), countries that had a universal BCG vaccination policy in the past (in green) and countries that currently have a universal BCG vaccination policy (in blue). 78 countries with information on BCG vaccination policy and at least 500 COVID-19 cases as of April 7^th^, 2020 are represented. **(B)** Total COVID-19 cases shown as mean ± SEM for all three groups of BCG policies (never (red), past (green) and current (blue) BCG vaccination policy) for countries with at least 500 confirmed COVID-19 cases. **(C**,**D)** Total COVID-19 cases per 1M inhabitants shown as mean ± SEM **(C)** or individual values **(D)** for all three groups of BCG policies. **(E**,**F)** Percent mortality per total COVID-19 cases shown as mean ± SEM **(E)** or individual values **(F)** for all three groups of BCG policies. Multivariate analysis has been performed, not significant (ns), p ≥ 0.05, * p < 0.05, ** p < 0.01, **** p < 0.0001.

Next, the following parameters were assessed in the univariate analysis: (a) population density (population/km^2^), (b) sex (ratio of female and male inhabitants), (c) heart-associated mortality, (d) percent smokers, (e) percent urban population, (f) percent migrants, (g) age, (h) rate of CoV-2 diagnostic testing, and treating a nation’s BCG vaccination policy as a monotonically increasing continuous variable (never BCG = 0, prior BCG = 1, current BCG = 2). Univariate regression analysis on all nations showed that percent urban population, percent migrants, age, and rates of testing confound the analysis (**Figure 2A**). The correlation between specific CoV-2 testing and BCG policy was robust (Pearson ρ = 0.82, p = 2.4×10^×19^, **Figure 2B**). A similar strong, but negative relationship, was observed between percent mortality and testing rates (ρ = -0.42 p = 1.5×10^×4^, **Figure 2C**). CoV-2 testing rates were significantly different between countries with distinct BCG vaccination policies (one-way ANOVA p = 6.5×10^×5^). The non-overlapping values between different countries with different BCG policies preclude robust multivariate analysis. To account for this in the subsequent analysis, we only included “high CoV-2 testing” countries with 2,500 or more tests per million (N: Current universal BCG vaccination policy = 21, universal BCG vaccination policy in the past = 17, Never universal BCG vaccination policy = 6), resulting in equivalent distribution of CoV-2 testing rates across countries with different BCG policies (one-way ANOVA p = 0.17). When comparing countries with high CoV-2 testing, current universal BCG policy is no longer significantly associated with a reduced number of COV-19 cases per million (**Figure 2D**). The number of COVID-19 cases per million was significantly lower in countries with a current universal BCG policy and low CoV-2 testing rates (**Figure 2D**). For mortality rates in high CoV-2 testing countries, there was a significant reduction in countries with current BCG vaccination policies, but no difference was observed in countries with current BCG vaccination policies and low CoV-2 testing rates (**Figure 2E**). Univariate regression analysis for number of COVID-19 cases per million in high-testing nations indicated that testing rate, percent heart deaths, and urban population remained as significant co-variates (**Figure 2F**), though none were significantly different based on a nation’s vaccination policy (one-way ANOVA p values of 0.17, 0.14, and 0.27, respectively). Univariate regression of percent mortality in high testing nations did not reveal any significant co-variates (**Figure 2G**). The distinct suppliers for the BCG vaccines did not correlate with number of COVID-19 cases and mortality, largely because of the scarcity in available data (**Figure 2H**).

**Figure 2.**
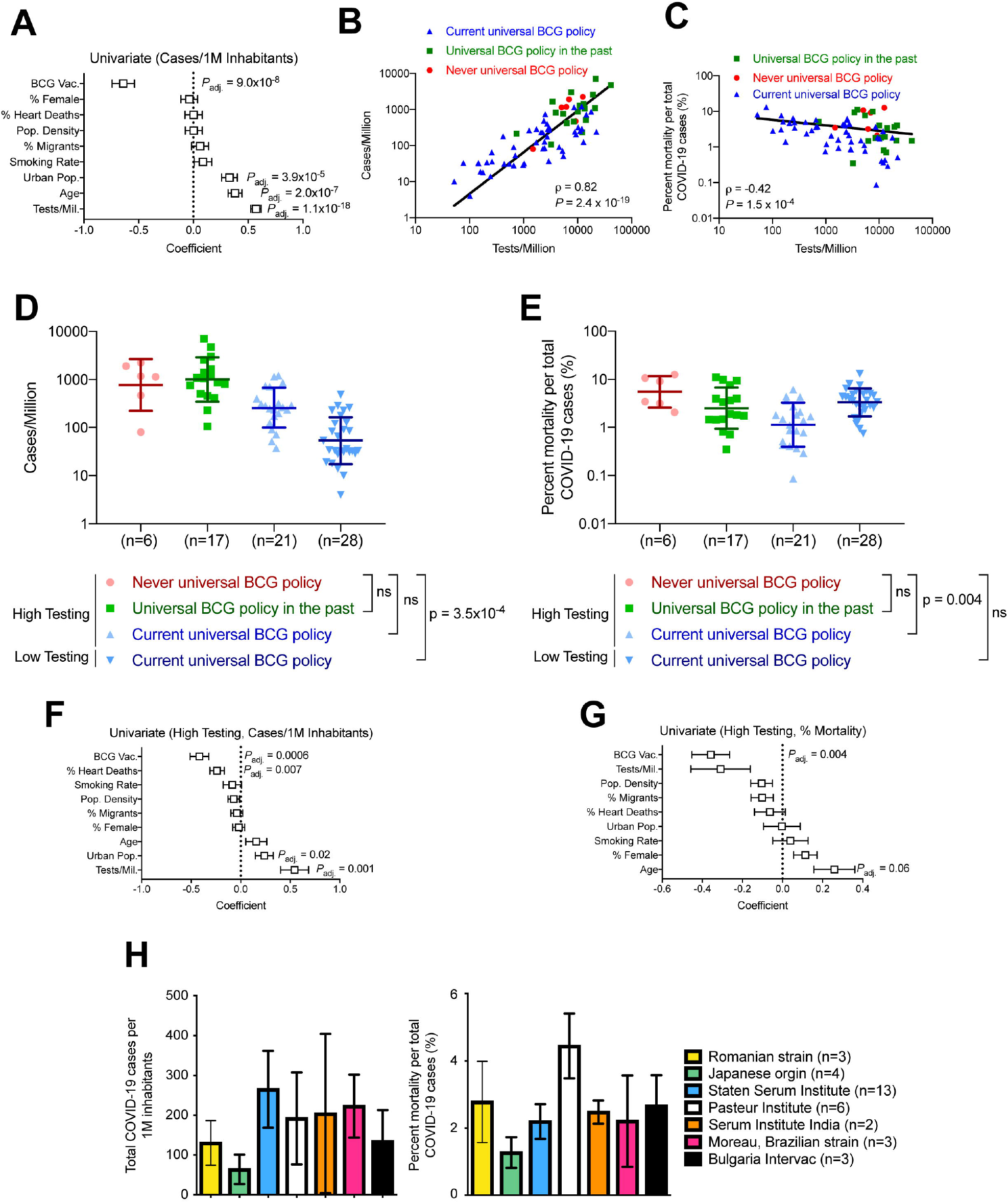
SARS-CoV-2 testing rates influence the observed benefit of BCG vaccination policy. **(A)** Univariate analysis of total COVID-19 cases per 1M inhabitants shows the coefficients and adjusted p-values for BCG vaccination policy (BCG Vac.), population density (pop. density), percent heart-related mortality (% heart deaths), smoking rate, urban population (urban pop.), percent migrants, age and test availability/1M inhabitants (tests/mil.). **(B)** Correlation graph of total COVID-19 cases per 1M and tests per 1M inhabitants. Inset value is Pearson’s correlation coefficient (ρ). Kruskal-Wallis with Dunn’s post-hoc test performed. **(C)** Correlation graph of percent mortality per total COVID-19 cases (death rate) and tests per 1M inhabitants. Inset value is Pearson’s correlation coefficient (ρ). Kruskal-Wallis with Dunn’s post-hoc test performed. **(D)** COVID-19 cases per 1M inhabitants is shown for all three groups of BCG policies (never (red), past (green) and current (blue) BCG vaccination policy), split by testing rates. Data shown as geometric mean ± geometric S.D. **(E)** Percent mortality per COVID-19 cases is shown for all three groups of BCG policies (never (red), past (green) and current (blue) BCG vaccination policy), split by testing rates. Data shown as geometric mean ± geometric S.D. **(F)** Univariate analysis of high testing countries (>2,500 tests per 1M inhabitants) for total COVID-19 cases per 1M inhabitants. **(G)** Univariate analysis of high testing countries (>2,500 tests per 1M inhabitants) for mean percent mortality per total COVID-19 cases (death rate). **(H)** Mean ± SEM of total numbers of COVID-19 cases per 1M inhabitants (left panel) and percent mortality per total COVID-19 cases (right panel) was plotted for different BCG strains in countries that currently have a universal BCG vaccination policy. Not significant (ns).

Our analyses also confirmed the association with implementing universal BCG policy with countries with high TB incidence (**Figure 3A**). In the countries with universal BCG policy in the past, the year in which the policy was discontinued was available for 14 out of the 18 countries in this group. The number of COVID-19 cases per 1M inhabitants was not significantly different when comparing countries that had discontinued the universal policy for at least 20 years and countries that had discontinued the universal policy less than 20 years ago (**Figure 3B**). The mortality associated with COVID-19 was not impacted whether the policy for universal BCG vaccine was discontinued less than or more than 20 years ago (**Figure 3B**). In the countries with current universal BCG policy and with available data, there was no correlation observed in the total COVID-19 cases per 1M inhabitant or percent mortality per total COVID-19 cases, and the time since implementation of the universal BCG policy (**Figure 3C**). There were also no significant correlation observed in the percent mortality per total COVID-19 cases and the time since the first confirmed COVID-19 cases in countries that never had a universal BCG policy (**Figure 3D**), or had a universal BCG policy in the past (**Figure 3E**), or have a current universal BCG policy (**Figure 3F**).

**Figure 3.**
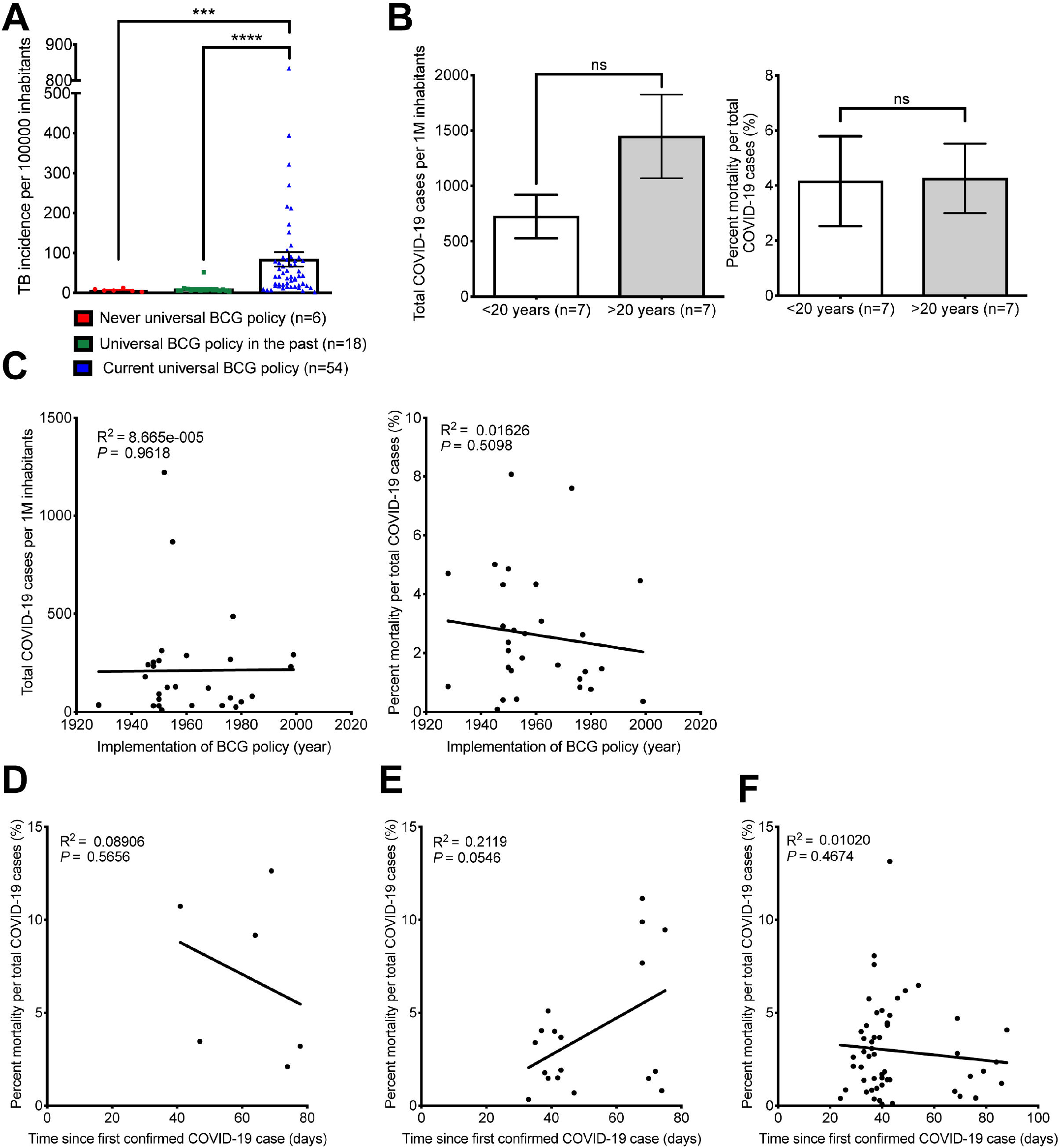
Distribution of COVID-19 cases and mortality and BCG vaccination policy implementation. **(A)** TB incidence in per 100,000 inhabitants shown as mean ± SEM for all three groups of BCG policies (never (red), past (green) and current (blue) BCG vaccination policy). One-way ANOVA with Kruskal-Wallis performed. **(B)** Total COVID-19 cases per 1M inhabitants (left panel) and percent mortality per total COVID-19 cases (right panel) is shown as mean ± SEM for all countries that had a universal BCG policy in the past and discontinued that policy ≤ 20 years ago or ≥ 20 years ago. Mann-Whitney test performed. **(C)** Correlation graph of total numbers of COVID-19 cases per 1M inhabitants (left panel) percent mortality per total COVID-19 cases (right panel) and the year the universal BCG policy was introduced. **(D**,**E**,**F)** Correlation graph showing percent mortality per total COVID-19 cases and the time since first confirmed COVID-19 case in days in countries that never had a universal BCG policy **(D)**, had a universal BCG policy in the past **(E)**, or have a current universal BCG policy **(F)**. Not significant (ns), p ≥0.05. *** p < 0.001, **** p <0.0001.

## DISCUSSION

Our independent analysis of COVID-19 cases per million of inhabitant in 78 countries that were stratified based on the prevalence of BCG vaccination (universal vaccination policy) shows that, a priori, there is a correlation between current universal BCG vaccination policy and a lower incidence of CoV-2 infection and related deaths. However, we note that testing rates (tests per million inhabitants) was positively correlated with incidence of CoV-2 infection. Taking in consideration this important bias, we curated the data to compare the number cases amongst countries with distinct universal BCG policy based on high CoV-2 testing rates, as defined by 2,500 tests or more per million inhabitants. The results show a lack of significant difference in incidence of COVID-19 cases in countries with current universal BCG vaccination policy and countries with never- or in the past-universal BCG vaccination policy. Other variables tested included population density, percent urban population, percent migrants, median age, sex, time since the first confirmed COVID-19 case, percent smokers, and heart associated mortality. Urban population and median age were significantly associated with increased number of cases; however, their impact were not as significant as CoV-2 testing rates. Interestingly, COVID-19 associated mortality was decreased in countries with current universal BCG vaccination policy and similar CoV-2 testing rates. This may however reflect potential inaccuracies in reporting of COVID-19 mortalities due to lack of Cov-2 testing. Our study is limited by several critical considerations, which were not addressed here. These include the variability and inaccuracies in reporting positive cases and reporting specific mortality associated with CoV-2 infection, lack of reporting on asymptomatic or minimally symptomatic cases, the impact of social and economic barriers in accessing care, the association with other co-morbidities, and the adherence or access to vaccination as per policy. In this regard, a recent report showed a correlation of universal BCG vaccination policies and the number of COVID-19 cases when controlling for income [7]. This study however did not take in consideration CoV-2 testing rates. It is also possible that the lower rate of CoV-2 testing, combined with lower viral loads in traditionally warmer countries with higher incidence of tuberculosis, could impact our observations [8]. Although not studied here, host haplotypes may also inform on the incidence COVID-19 cases. Finally, time since first exposure rates, as well as population adherence to social distancing measures, may also influence the evolving COVID-19 data collection. To date, social distancing remains critical in slowing down the number of new cases over time. Data from the Australian clinical testing currently underway to assess whether the BCG vaccination may be protective against COVID-19 (clinicaltrials.gov, NCT04327206) will likely shed more light on the possible use of BCG in decreasing CoV-2 infection and associated mortality.

## Data Availability

Data are publicly available.

